# Declines in HIV Incidence and Prevalence and Predictors Among Adolescents and Young Adults: An Observational Cohort Study, Rakai, Uganda, 2005-2020

**DOI:** 10.1101/2025.01.07.25320118

**Authors:** Stephanie A Grilo, Julia Thompson, Ivy S Chen, Fred Nalugoda, Tom Lutalo, Ying Wei, Esther Spindler, Susie Hoffman, Philip Kreniske, David Serwadda, Joseph Kagaayi, Mary Kate Grabowski, Maria J Wawer, Fred M Ssewamala, Larry W Chang, Aleya Khalifa, Debbie Malden, John S Santelli

## Abstract

**Background:** HIV acquisition among adolescents and young adults (AYA, 15-24 years) is influenced by individual factors, community factors, and public policies and programs. We explored the association of HIV incidence and prevalence with these factors over time among AYA in Rakai, Uganda.

**Methods:** We examined trends over nine survey rounds (2005-2020) of the Rakai Community Cohort Study (RCCS), an open population-based surveillance cohort of individuals living in 30 continuously followed communities in south-central Uganda (n= 35,938 person rounds). We evaluated the associations between individual and community-level factors including HIV community viremia (CV, a measure of community-level ART use and HIV prevalence) and HIV incidence and prevalence. Logistic GEE, Poisson GLM and univariate models were run for HIV prevalence, HIV incidence, and predictors of interest, respectively.

**Findings:** HIV incidence and prevalence declined over time after round 14 (2010-2011) by 66% among AYA men and after round 17 (2015-16) by 60% among young women. Between survey round 11 (2007-2008) and round 19 (2017-2019), the proportions reporting being sexually experienced declined from 58% to 38% in adolescent men (15-19) and from 65% to 35% among adolescent women. The prevalence of VMMC among adolescent men increased from 20% in round 11 to 79% in round 19. At the community-level, we found substantial increases in ART use among PLHIV ( 5% in round 11 and 86% in round 19) with corresponding declines in community viremia. In multivariable analyses, a combination of individual and community-level factors were found to predict HIV incidence and prevalence among AYA, notably VMMC among young men and community viremia among young women.

**Interpretation:** Declines in HIV incidence and prevalence occurred first among AYA men and later among AYA women. These coincided with declines in sexual experience and with public policies that increased access to VMMC and ART. Combination HIV prevention with AYA needs to address risk factors at multiple levels.

**Funding:** This work was supported by the Eunice Kennedy Shriver National Institute of Child Health and Human Development (NICHD, grants R01HD091003, RO1HD070769, R01HD050180, R01 HD074949, and P2CHD058486), the National Institute of Allergy and Infectious Diseases (grants R01AI143333, R01AI110324, U01AI100031, U01AI075115, R01AI110324, R01AI102939, and K01AI125086-01), the National Institute of Mental Health (grants R01MH115799, R01MH107275, R01 MH128232, F31MH134699), and the Division of Intramural Research of the National Institute for Allergy and Infectious Diseases.

**Author Contributions:** SAG, JSS, ISC, TL, YW, ES, SH, AND PK conceived and designed the study. ISC, JT and YW oversaw data cleaning and statistical analysis and directly accessed and verified the underlying data reported in the manuscript. AK and DM accessed the data, created the figures for the resubmission and edited the text. AK and DTL, FN, DS, JK, MKG, MJW, LWC, FMS oversaw data collection. All authors had full access to the data in the study, participated in the interpretation of data and revising the manuscript, and had final responsibility for the decision to submit for publication.

**Research in context:** *Evidence before this study:* Age-specific rates of HIV incidence are often highest among AYA and particularly young women. Risk factors for HIV infection among AYA include earlier sexual initiation, multiple partners, and inconsistent condom use. Combination prevention including community-wide uptake of ART and male medical circumcision has been associated with declines in HIV incidence in Rakai, Uganda in the overall population.^1,2^ A 2019 review of HIV incidence among adolescent girls and young women from 10 high-prevalence African countries found that few studies have examined incidence over time, and among those that there was limited evidence of incidence declines.^3^

*Added value of this study:* Using data from 2005-2020 and the Rakai Community Cohort Study (RCCS) in southcentral Uganda, we found evidence that community-level factors including community viremia and ART use, VMMC among young men, and declines in sexual experience were associated with lower risk of HIV acquisition and seroprevalent infection among AYA. Declining HIV incidence and prevalence over time among AYA coincided with policy changes expanding access to ART and VMMC.

*Implications of all the available evidence:* Age of sexual initiation and community-level factors play critical roles in HIV transmission in Rakai and in declines over time in youth HIV incidence and prevalence. HIV prevention for AYA needs to address individual factors and public policies to improve access to ART and VMMC.

## INTRODUCTION

Globally, HIV incidence and prevalence are highest in sub–Saharan Africa (SSA) with adolescent and young adults (AYA) at among the highest risk.^4,5^ Adolescent and young adult women (AYAW) accounted for 25% of new HIV infections in 2020, despite representing just 10% of the population.^4^ AYAW, 15-24 years are also at much higher risk compared to adolescent and young adult men (AYAM).

HIV prevention programs and policies should be guided by a nuanced understanding of HIV risk factors and how changes in risk factors and interventions over time may contribute to trends in HIV infection.^6,7^ Risk factors for HIV incident infection among AYA include individual biological and behavioral factors, social conditions, community-level factors including public policies and programs, and HIV prevalence within specific communities.^7^ Certain policies and programs protect individuals but may have community-wide impact; these include access to antiretroviral therapy (ART), voluntary male medical circumcision (VMMC), and PrEP.^1,8–10^ Among AYA, HIV infection may be influenced by social transitions such as leaving school, initiating sexual intercourse, marriage, and initiation of childbearing.^11^ Social determinants of HIV infection include poverty, gender inequality, stigma, and public policies that promote access to education, sex education, and health services.^7,12^ School enrollment and higher educational attainment are key protective factors among AYA in preventing new HIV infection.^7,13^ Programs like the DREAMS initiative have focused on social and behavioral determinants of HIV transmission^14^

Much of the population level data on risk factors is cross-sectional and does not explain changes over time in HIV infection and doesn’t include young men. Incidence is difficult to measure as it generally requires longitudinal data collection. Even well characterized risk and protective factors may not explain change over time, <u>if</u> those factors are not changing. For example, behavioral factors such as condom use or biological factors such as cervical ectopy increase infection risk but may show little change over time in a population. Moreover, considerable change in a risk factor may be needed to demonstrate change in HIV incidence within a population. Thus, countries may fail to detect declines in HIV incidence among AYA. A 2019 review by Birdthistle and colleagues of HIV incidence among adolescent girls and young women from ten high-prevalence African countries found evidence of declining incidence in only two cohorts (Manicaland, Zimbabwe and Rakai, Uganda).^3^ They found little evidence to suggest ART availability or other intervention efforts had slowed HIV transmission through 2016.

In Rakai, Uganda, HIV incidence among reproductive-age persons (15-49 years) declined after the scaling up of combination HIV prevention (CHP) including access to ART beginning in 2004 and VMMC in 2007.^1^ Our prior analyses among AYA in Rakai (1999-2011 and 1994-2013) demonstrated that the decline in HIV incidence was associated with improvement in individual behavioral risk factors (declines in sexual experience, multiple partners, and sexual concurrency) and social factors (increases in school enrollment, availability of ART and VMMC, rising SES, declining orphanhood.^1,2,11,13^ Much of the decline in HIV incidence among adolescent women was attributable to increased school enrollment and delayed initiation of sexual intercourse following Uganda’s 1997 national policy of universal primary education (UPE).^13^

This analysis extends our previous work in Rakai by quantitatively examining the relationship between community-level factors including ART use and VMMC and HIV incidence and prevalence and includes three additional survey rounds gathered over six years since our last analysis. We include both incidence and prevalence in the study because while HIV incidence reflects recent infections, it is difficult to measure, and most risk factor studies of HIV Infection have used prevalence. Thus, our prevalence analysis provides comparable data to other studies. The incidence analysis has a smaller sample and less power. Calculating both incidence and prevalence offers concordance in our findings. Our prior research comparing predictors of prevalent and incident HIV infection found that they are similar but not identical.^15^ We created a variable for “community viremia” which adjusts HIV prevalence in a community for the percentage of PLHIV on ART. Previous research on adolescents in Rakai in SSTAR did not examine community level factors and did not include the most recent year of data from the cohort.

## Methods

### Study Design, Procedures, and Population

We used data from 30 communities in rounds 11-19 (2005-2020) of the Rakai Community Cohort Study (RCCS), a population-based open cohort of individuals living in the Rakai District in south-central Uganda (n= 35,938 person rounds). Community-wide HIV education, individual and couple’s HIV counseling and testing, referral for voluntary male medical circumcision (VMMC), and antiretroviral therapy (ART) are offered free of charge.^1^ Currently, adolescents <15 years are not eligible to participate in the RCCS but are enumerated in the household census. Per Uganda’s National Clinical guidelines, both written minor assent and parental/guardian permission are obtained for unemancipated minors (<18 years); 18+ year-olds and emancipated minors provide their own written informed consent. Emancipated minors are legally considered adults or have been granted independence from parental control - in this sample they were interviewed and are either household heads, or married, or have a living child. For context, in this sample in the last two survey rounds (rounds 19 and 20) the percentage of young people who were considered emancipated minors was right around 5%.

Each round of the RCCS takes 1-2 years and consists of a household census, enrollment and verification, individual survey interviews, and collection of blood for HIV testing.^1^ All households within RCCS communities are included in the census, with data provided by the head of household. All individuals between the ages of 15 and 49 years within a household are eligible for an interview. Two to four weeks after the census, consenting residents (including both new members and those followed-up) are enrolled and asked to provide blood for HIV and sexually transmitted infection testing, treatment, and referral. Enrolled individuals are then interviewed extensively about sexual and reproductive health and HIV risk.

Data are entered electronically in the field and are immediately reviewed by supervisors/editors to ensure data integrity. RCCS participation rates are about 95% of those present at time of survey; about 25% of all censused residents are absent at each round. Acceptance of HIV testing among enrollees is high (>95%).

We used data from 30 RHSP communities which have been surveyed continuously over time (every survey round) from 2005 to 2020 (rounds 11-19). Our analyses focus on AYA age 15 to 24 years at the time of the survey. Data were selected to begin at round 11 (2005-2006) because key questions for our analysis were not asked consistently in earlier survey rounds. We note that round 19 (2018-2020) included data collection both before and during the COVID-19 pandemic.

Approvals were obtained from Uganda Virus Research Institute’s REC, Uganda National Council for Science and Technology, and IRBs at Columbia University and Johns Hopkins University.

### Variables

Data came from several sources described above including: the census, the RCCS questionnaire, and HIV testing. We used a combination of individual-level variables (age, marital status, VMMC, ART use, school enrollment, sexual initiation, condom use, and number of sex partners) and community-level variables (HIV prevalence, community viremia).

#### Individual-level variables

Participant characteristics and other HIV risk factors (age, marital status, male medical circumcision (VMMC), ART use among people living with HIV, school enrollment, sexual initiation, condom use, and number of sex partners) were obtained through self-report from the survey questionnaire. Age is asked in the interview and is double-checked using the census. Participants are asked if they have ever been married and if they are currently married. Men are asked if they are circumcised, and ART use is asked if a person reports being HIV positive. Self-report of ART use has been validated previously with a specificity of 99% and a sensitivity of 77%.^16^ A participant is considered to be enrolled in school if they answer “student” to the occupation question on the survey.^11^ Educational attainment comes from asking the participant if they have ever attended school and to what level. The RCCS asks about the four most recent sexual partners and examines behaviors such as condom use within each partnership. Consistent condom use in the past 12 months is defined as answering “always” with all partners. The household socioeconomic status (SES) measure was derived from the census using principal component analysis. The census collects nine household assets (e.g., ownership of a bicycle, car, motorcycle) including home construction (e.g., metal roof). SES was standardized based on the distribution of scores over 19 rounds of RCCS data collection (1994-2018) using a z-transformation.

HIV infection was obtained through testing provided during the RCCS survey interview. HIV incidence analyses included participants who had at least two rounds of data and skipped no more than one round in between.^1^ An incident case was a person who was HIV negative at the first round and then was HIV positive at the next round. Individuals who were HIV-positive at enrollment or had more than one intermediate round missing were removed from the incidence analysis.

#### Community-level variables

People living with HIV who adhere to ART are much less likely to transmit HIV to others (Saag et al., 2019). By effectively blocking HIV replication, ART helps to reduce the viral load in PLWH, contributing to improved health outcomes and decreased transmission risk. Community viremia is the community HIV prevalence adjusted for the proportion of individuals within a community who have initiated ART. Community viremia was defined as community-level HIV prevalence minus 90% of the proportion of those individuals living with HIV who report being on ART, i.e.,: Viremia=HIV Prevalence-(90%*ART Coverage*HIV Prevalence).^1^ To estimate HIV risk among AGYW, we calculated community viremia based on data from men age 15-34 years and to estimate risk among ABYM we calculated community viremia based on data from women age 15-30 years. These age ranges were selected as they are the age groups most likely to transmit HIV to AYA.^17^ AYA women have somewhat older male partners up to approximately age 34, while AYA men have slightly younger female partners up to approximately age 30.^11,18^ These variables were computed at the community level, and each community was assigned a value for each round.

### Analysis

The analysis was performed using R version 4.2.1 or later. Each factor of interest (participant characteristics, and HIV prevalence and incidence) was summarized by round and age group. Participant characteristics were examined separately by gender. Linear trends were fit for all factors to examine change over time. Quadratic trends for time were also fit for participant characteristics. Linear and quadratic provide different information. Over long periods of time, the direction of change over time may change. Quadratic (often curvilinear) suggests an inflection point in slope or non-linear change over time. A conceptual model) was created to conceptualize the relationships between variables and decide which variables should be included in the modeling (see Figure 1 in supplementary materials page 3). Models were selected according to the variable type. Logistic generalized estimating equations (GEE) was used for dichotomous outcomes, and Gaussian GEE for continuous outcomes. Multinomial or ordinal logistic regression with robust standard errors was used for multivariate outcomes (HIV prevalence). Poisson Generalized Linear Models (GLM) were used for HIV incidence.

**Figure 1.**
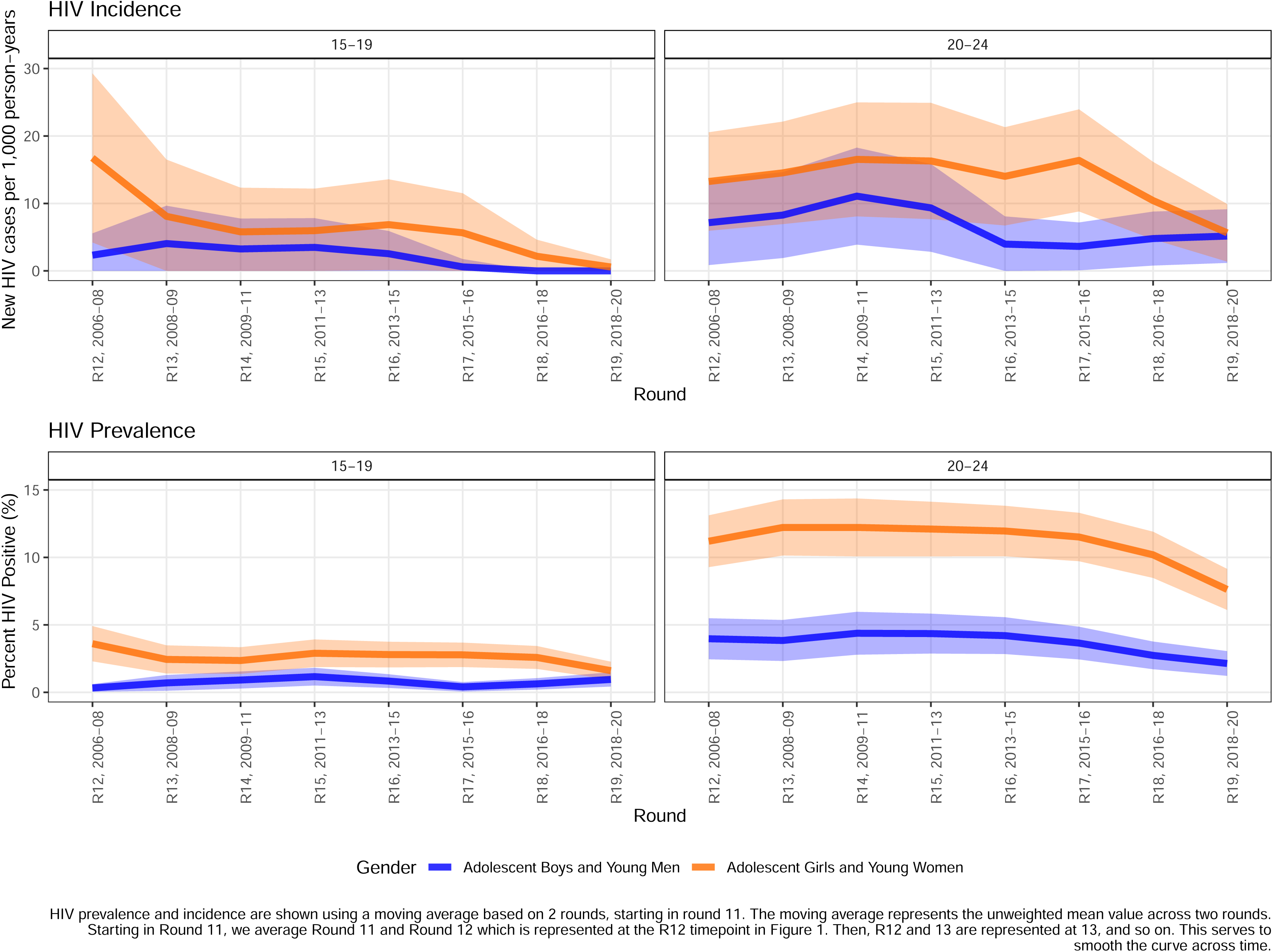
Trends in HIV among Adolescents and Young People in 30 Communities, Rakai District, Uganda, 2005−2020 HIV Incidence.

Our primary outcomes were HIV seroprevalent infection and HIV seroincident infection. As described above, logistic GEE models were run for HIV prevalence and Poisson GLM were run for HIV incidence, separately by gender. Predictors were informed by knowledge of HIV prevalence and incidence among AYA, and included round, marriage status, number of sex partners, SES category, age group, community viremia, and VMMC for men.

## Results

The data include 9 survey rounds, with an average of 2,164 young women and 1,834 young men participating per round. There are a total of 19,473 and 16,510 person-rounds of data for young women and young men, respectively.

HIV incidence declined after round 14 (2010-2011) among AYA men and after round 17 (2015-2016) among young women (Figure 1 and supplemental table 1 page 4). HIV incidence declined 60% among AYA women from round 17 (2015-2016) to round 19 (2018-2020) (12·1 to 4·9 per thousand, p=0·09) and 66% among AYA men from round 14 (2010-2011) to round 19 (2018-2020) (8·9 to 3·0 per thousand, p=0·07). The timing of declines in HIV prevalence had a similar pattern - declining first among young men and then among young women (Figure 1, Supplemental Table 1 page 4). Among young women, the decline was 51% from round 17 (2015-2016) to round 19 (2018-2020) (7·1% to 3·5%, p<0·001). Among young men, the decline in prevalence was 44% from round 14 (2010-2011) to round 19 (2018-2020) (2·5% to 1·4%, p=0·03). For young adult women, HIV prevalence increased over time and peaked in round 13 (2008-2009) before declining again (supplementary table 1 page 4, p<0.001). HIV incidence and prevalence were consistently higher among AYA women compared to AYA men and higher among 20-24-years-old compared to 15–19-year-olds.

Social and Behavioral HIV Risk Factors (Figures 2 and 3 and supplemental Table 2 and 3 pages 5-9)

**Figure 2.**
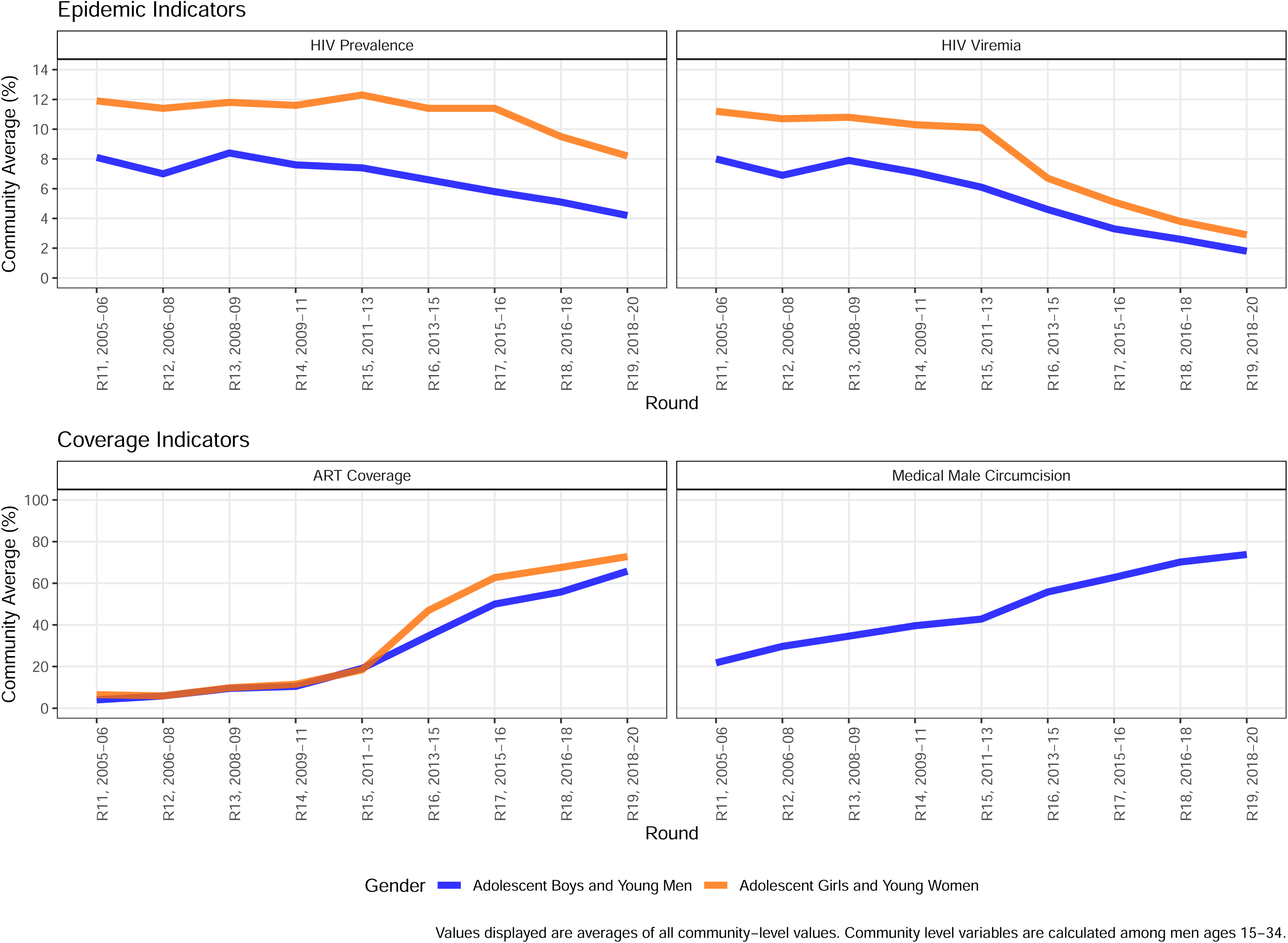
Trends of Community−level HIV Indicators among Adolescents and Young People in 30 Communities, Rakai District, Uganda, 2005−2020.

**Figure 3.**
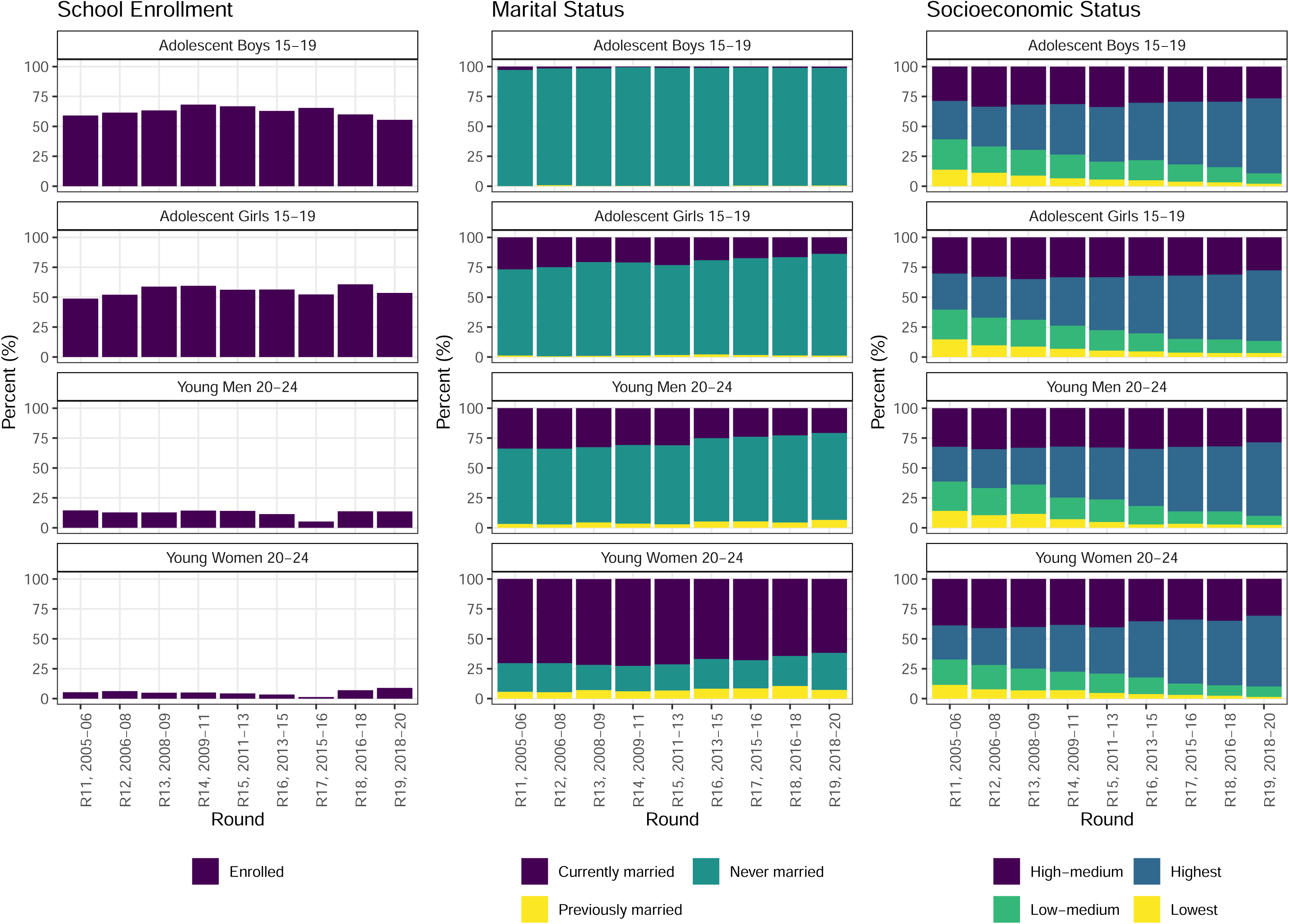
Trends in Social HIV Risk Factors among Adolescents and Young People in 30 Communities, Rakai District, Uganda, 2005−2020.

Among women, school enrollment increased to round 14 (2010-2011), plateaued and then declined in round 19 (2018-2020) among both adolescents and young adults (quadratic trends, p<0·001 and p<0·001). Marriage (ever and current) decreased over time among both adolescent and young adult women. Household SES increased steadily over time, as we have previously reported.^19, 20^

Reporting ever having had sex decreased among adolescents (p<0.001), from 65% in round 11 (2005-2006) to 35% in round 19 (2018-2020); ever sex also declined among young adults (98% in round 11 (2005-2006) to 90% in round 19 (2018-2020)). However, among those sexually experienced, having two or more sexual partners increased among both adolescent (10% to 14%, p<0.001) and young adult women (8% to 12%, p<0.001).Consistent condom use with most recent partner declined from 25% to 17% (p<0.001) among adolescents and from 9% to 5% (p=0.001) among young adults.

Among adolescent men, trends in school enrollment were similar to those among adolescent women; enrollment increased through round 14 (2010-2011) and then decreased (linear and quadratic trends (p<0·001, Table 2). Marriage was not common among adolescent men; among young adult men, the proportion who were never married increased from 63% to 73% while that of those currently married decreased from 34% to 21%. Household SES increased steadily over time.

Among men, reporting ever had sexual experience declined from 58% to 38% among adolescents and from 95% to 87% from young adult men. VMMC increased considerably over time across both age groups, from 20% in round 11 (2005-2006) to 79% at round 19 (2018-2020) among adolescents and from 22% to 72% among young adult men. Having two or more sex partners in the past year showed a quadratic trend among both adolescent and young adult men, decreasing to the lowest level at round 15 (2011-2013) and then increasing. Consistent condom use with most recent partner did not change for adolescents but decreased for young adult men from 34% to 25%. Condom use with 2-4^th^ partners showed no change over time.

Community Level HIV Risk Factors (Figure 4 and supplemental table 4 page 10)

**Figure 4.**
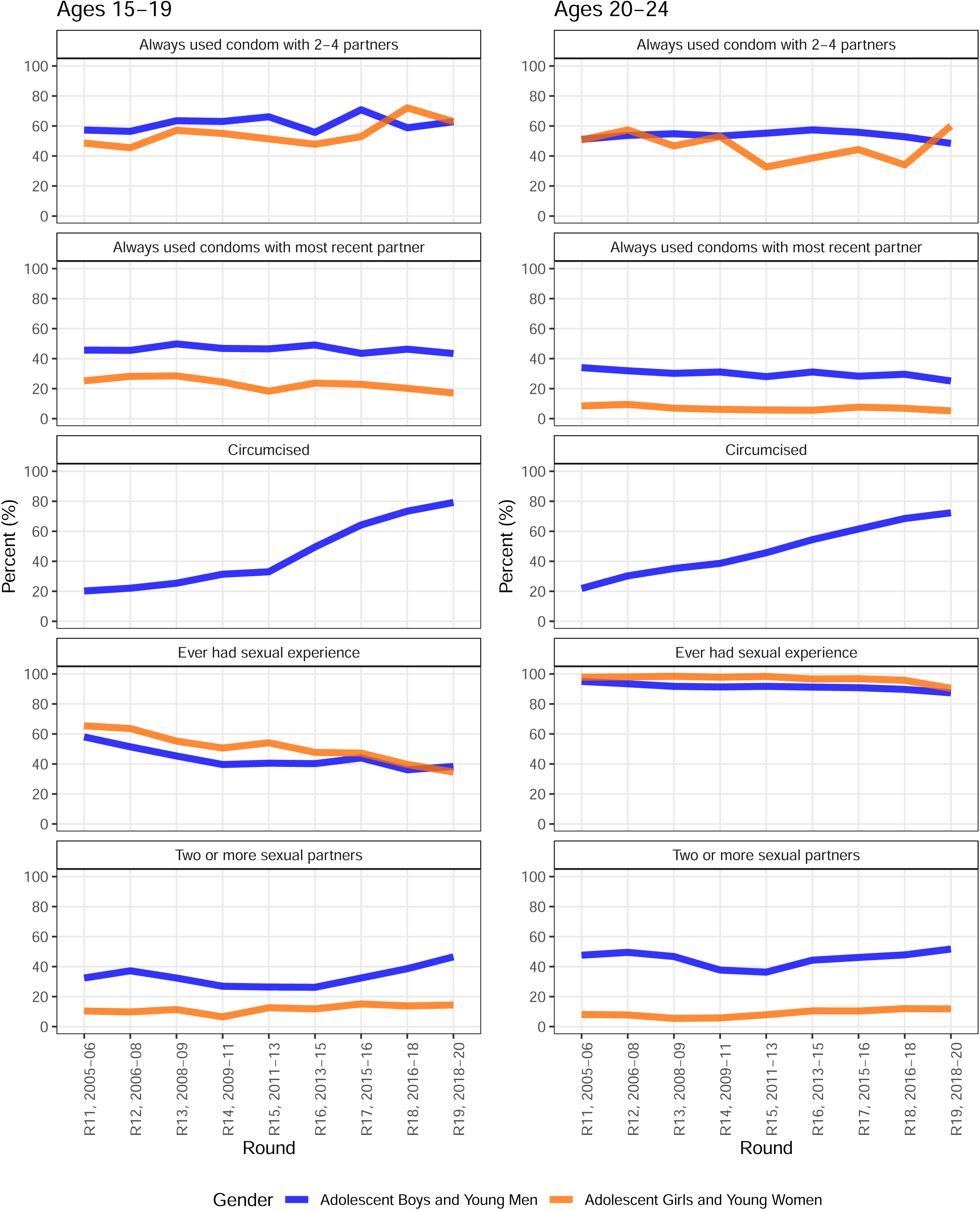
Trends in Behavioral HIV Risk Factors in 30 Communities, Rakai District, Uganda, 2005−2020.

Population-wide, ART use among PLHIV in Rakai rose from 5% in round 11 (2005-2006) to 86% in round 19 (2018-2020) (Supplemental Table 4 page 10), while HIV prevalence did not change over the same time period. Community viremia which is based on both ART use and HIV declined by 70%, from 14·6% to 4·4%. The population prevalence of MCC rose from 24% in round 11 (2005-2006) to 66% in round 19 (2018-2020). Trends in VMMC accelerated over time among adolescent men (quadratic trend, p<0·001); this pattern corresponds to both the randomized control trial of VMMC in Rakai (2003-2007) and to the roll out of VMMC as a routine service after 2007.

Supplementary figure 2 (page 13) displays the univariate associations between selected individual- and community-level factors and HIV prevalence among AYA women and men. In univariate models, being previously married and older were the strongest risk factors for HIV prevalence. Being previously married was associated with 5.44 (95% CI: 3.65-8.13) and 3.36 (95% CI: 2.61-4.34) greater odds of prevalent HIV infection compared to never having been married among AYA men and women, respectively. Compared to 15–19-year-olds, persons aged 20-24 years had approximately 4-fold greater odds of prevalent HIV infection among AYA men (OR: 4.04, 95% CI: 2.88-5.64) and 2-fold greater odds of prevalent HIV infection among AYA women (OR: 2.01, 95% CI: 1.81-2.23), respectively. Multiple sexual partners in the previous year, and increased community viremia were also associated with higher risk of prevalent HIV infection compared to 0-1 sexual partners and lower community viremia. Before adjustment, VMMC and high SES were associated with reduced HIV risk compared to no VMMC and lower SES categories. Although there was less statistical power to detect an association, in general, univariate associations were similar for HIV incidence (Supplementary figure 3 page 14).

In multivariable analyses, a combination of individual and community-level factors were found to predict HIV incidence and prevalence among AYA, notably VMMC among young men and community viremia among young women. **Figure 5** summarizes the associations between VMMC and community viremia and risk of HIV prevalence and incidence among AGYW and ABYM. Measures of association were adjusted for survey round, age group, marital status, household SES, school enrollment, and number of sexual partners. Full models are found in Supplemental Materials, Tables 5 and 6. Among ABYN, VMMC was associated with a 58% reduction in HIV incidence (IRR: 0.42, 95% CI: 0.23-0.76) and a 36% reduction in HIV prevalence (OR: 0.64, 95% CI: 0.48-0.86). Each 10% increase in community viremia was associated with 73% increased risk of HIV prevalent infection (OR: 1.73, 95% CI: 1.21-2.47) and 85% increased risk of HIV incident infection (IRR: 1.85, 95% CI: 0.90-3.81) among ABYM. Although the same increase in community viremia among AGYW was still associated with increased HIV prevalence (OR: 1.39, 95% CI: 1.17-1.65) and HIV incidence (IRR: 1.71, 95% CI: 1.03-2.84), associations were weaker than among AYA men, respectively. Risk estimates were generally similar and directionally concordant for both HIV prevalence and incidence.

**Figure 5.**
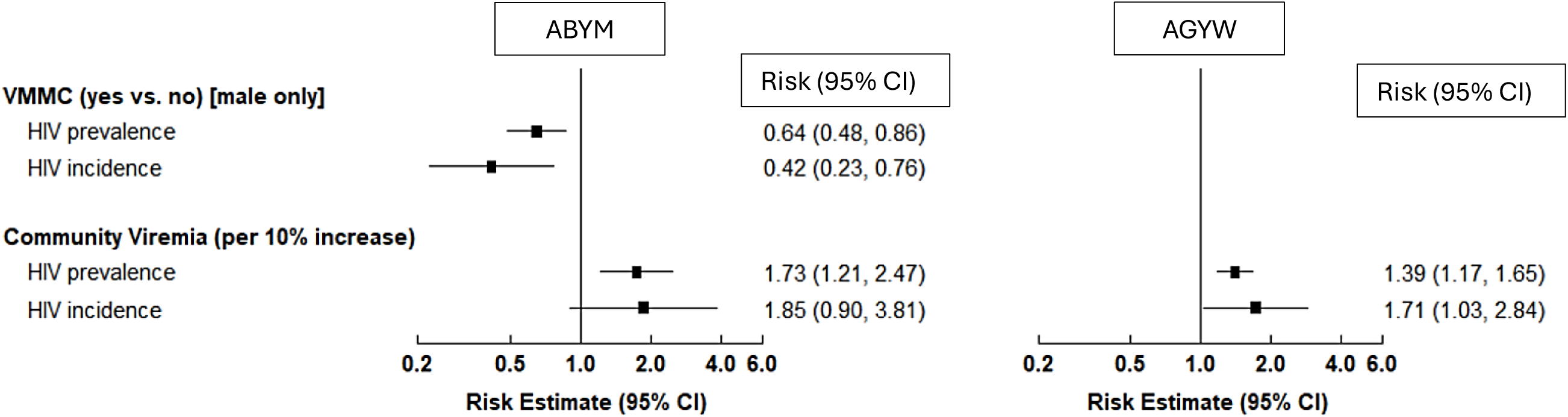
Comparison of community-level predictors for HIV prevalence and incidence among adolescents and young adults, Rakai District, Uganda, 2005-2020. *Multivariable models for HIV prevalence show adjusted odds ratios. Multivariable models for HIV incidence show adjusted incidence rate ratios. Each model adjusted for survey round, age group, marital status, household SES, school enrollment, number of sexual partners, community viremia and VMMC for men. Full models found in Supplemental Materials, Tables 5 and 6.

## Discussion

We found substantial declines in both HIV incidence and prevalence among AYA in Rakai - first among young men after 2011 and then among young women after 2016. The current findings align with World Bank estimates for Uganda, showing that AGYW have been disproportionately affected by the HIV epidemic, with consistently higher incidence and prevalence than ABYM. Both groups experienced declines in HIV prevalence and incidence over the study period, consistent with World Bank data.

Alongside these declines were community-wide increases in use of ART and VMMC and decreases in sexual experience among adolescents. Declines in HIV incidence and prevalence coincided with the implementation of public policies and resources to increase access to VMMC and ART.^1,2,21^

We identified a number of well-characterized risk factors for HIV incidence or prevalence which are unlikely to explain the declines in HIV incidence and prevalence in Rakai beginning after round 14 (2010-2011) for young men and after round 17 (2015-2016) for AYA women. Condom use (a protective factor) decreased over time and thus cannot explain a decrease in HIV incidence and prevalence.

Likewise, having multiple partners is a well described risk factor for HIV and other STIs. However, the prevalence of multiple partners among sexually experienced AYA increased over time - making it unlikely to explain decreases in HIV incidence or prevalence. Similarly, household socioeconomic status increased linearly from 2005 to 2020 and thus the trajectory of SES increases was not consistent with timing of the decline in incidence and prevalence - after rounds 14 (2010-2011) and 17 (2015-2016) for men and women, respectively.

While both individual and community-level factors were predictors of HIV incidence and prevalence most did not explain change over time. VMMC and ART use emerged as potential drivers of the decline in HIV infection among AYA in Rakai, warranting future causal analyses to estimate each of their effects on the youth epidemic. Both are well-established factors in preventing HIV transmission. The timing of the declines in HIV incidence and prevalence also follows closely upon the roll out of ART after 2004 within Rakai communities and the corresponding decline in community viremia. The pattern of increasing VMMC among AYA men (which began in 2005 and accelerated after 2009) corresponds to both the randomized controlled trial of VMMC in Rakai and to the roll out of VMMC as a routine service after 2007.^1,2,21^

VMMC directly protects men and - over time - may indirectly protect women by lowering prevalence among men.^21,22^ The earlier decline in HIV incidence and prevalence among AYA men compared to AYA women is consistent with this pattern of direct and indirect protection. These patterns of change in our data from AYA are similar to the declines reported among the entire Rakai cohort (men and women 15-49 years).^1,2^ Those analyses demonstrated that the incidence of HIV infection declined significantly following the scaling up of combination HIV prevention.^1^

The decline in sexual experience among Rakai adolescents was sizable and may have contributed to the decline in HIV incidence and prevalence for adolescent women and men, though more research is needed to determine its effect. It may have a much smaller impact among young adults as most had initiated sex by the early 20s. Delaying initiation of sex among adolescents can be effective in reducing HIV transmission.^23^

Explaining the decline in sexual experience is more complicated. In our prior research on HIV trends among Rakai adolescents through 2011, decline in sexual experience was highly associated with rising school enrollment.^13^ In these newer data, we note that school enrollment among adolescents plateaued after 2011 and declined in round 19 (2018-2020) during COVID-19 school closures. Similar patterns in school enrollment have been reported for Uganda since 2007.^24^ Thus, school enrollment may no longer be the primary driver of trends in sexual initiation in Rakai. Our updated analysis through 2020 strongly suggests a change in our understanding of HIV risk among AYA. The new data suggest that ART and VMMC may now be major drivers of the decline in HIV infection among AYA in Rakai. This is consistent with findings from other studies that have demonstrated the role of both community level as well as individual level risk factors for HIV incidence.^25^

An important contribution of this analysis is the inclusion of community-level risk factors for HIV incident and prevalent infections among AYA. The creation and inclusion of a community viremia variable, and the ability to look at VMMC at both the individual and community level allowed us to explore temporal trends of combination prevention and how they relate to trends of individual risk of HIV incidence and prevalence. Another contribution of this analysis is the modeling of both HIV incidence and prevalence. By examining both outcomes, we were able to understand epidemiologic trends more completely.

### Limitations

These analyses have several limitations. First, the majority of our measures, aside from HIV infection, were based on self-reported survey data. With this type of sensitive self-reported data, there may be a risk of social desirability bias and recall error. However, the RCCS, as one of the oldest and largest population-based studies in Southern Africa, has extensive procedures to assure validity and reliability.

For example, ART self-report use has high specificity and moderate sensitivity in this population.^16^ Likewise, participant-reported circumcision status has been validated previously from Rakai clinical records, with a specificity of 90%”.^21^ Second, because the RCCS is an open cohort, participants may move in and out of the study area and therefore the sample. Mobility is high, and in-migrants may differ meaningfully from out-migrants. Third, selection bias should be considered – participation is high (∼95%) in the RCCS of individuals present at the time of the survey, however about 25% are absent at a specific round, typically due to work or school. Fourth, the Rakai district is primarily rural and RCCS findings may not be generalizable to other settings. However, prior studies have found that the RCCS is representative of the broader Ugandan population.^26^ Finally, we do not have data to assess the impact of other interventions such as DREAMS on this population.

### Implications

This analysis suggests that individual risk behaviors may continue to play a role in HIV incident and prevalent infection. Thus, it remains important to have conversations with adolescents about their individual behaviors. However, community level factors may play an important role and therefore these conversations should occur within the context of larger social forces of transmission risk.

Thus, it is not sufficient to focus on individual behaviors to prevent HIV infection among AYA. HIV prevention with AYA needs to consider community access to ART and VMMC across the lifespan, and policies and programs that delay initiation of sex. More analysis is needed to understand the proportion of HIV risk among AYA attributable to each of these factors. Future research could employ causal inference methods and leverage longitudinal population-based data sources like the RCCS to determine causal effects.

## Data Availability

A deidentified version of the RCCS data may be provided to interested parties subject to completion of the
Rakai Health Sciences Program data request form and signing of a Data Transfer Agreement. Inquiries should be directed to.

## Acknowledgements

We thank the NIH for funding this research. We would like to thank the participants of the RCCS and the Rakai Health Sciences researchers for their invaluable contributions. We would also like to thank Kate Hicks and Meg Nolta for their assistance with the preparation of this manuscript.

## Data Sharing

A deidentified version of the RCCS data may be provided to interested parties subject to completion of the Rakai Health Sciences Program data request form and signing of a Data Transfer Agreement. Inquiries should be directed to.

---

**Supplemental Figure 1.**
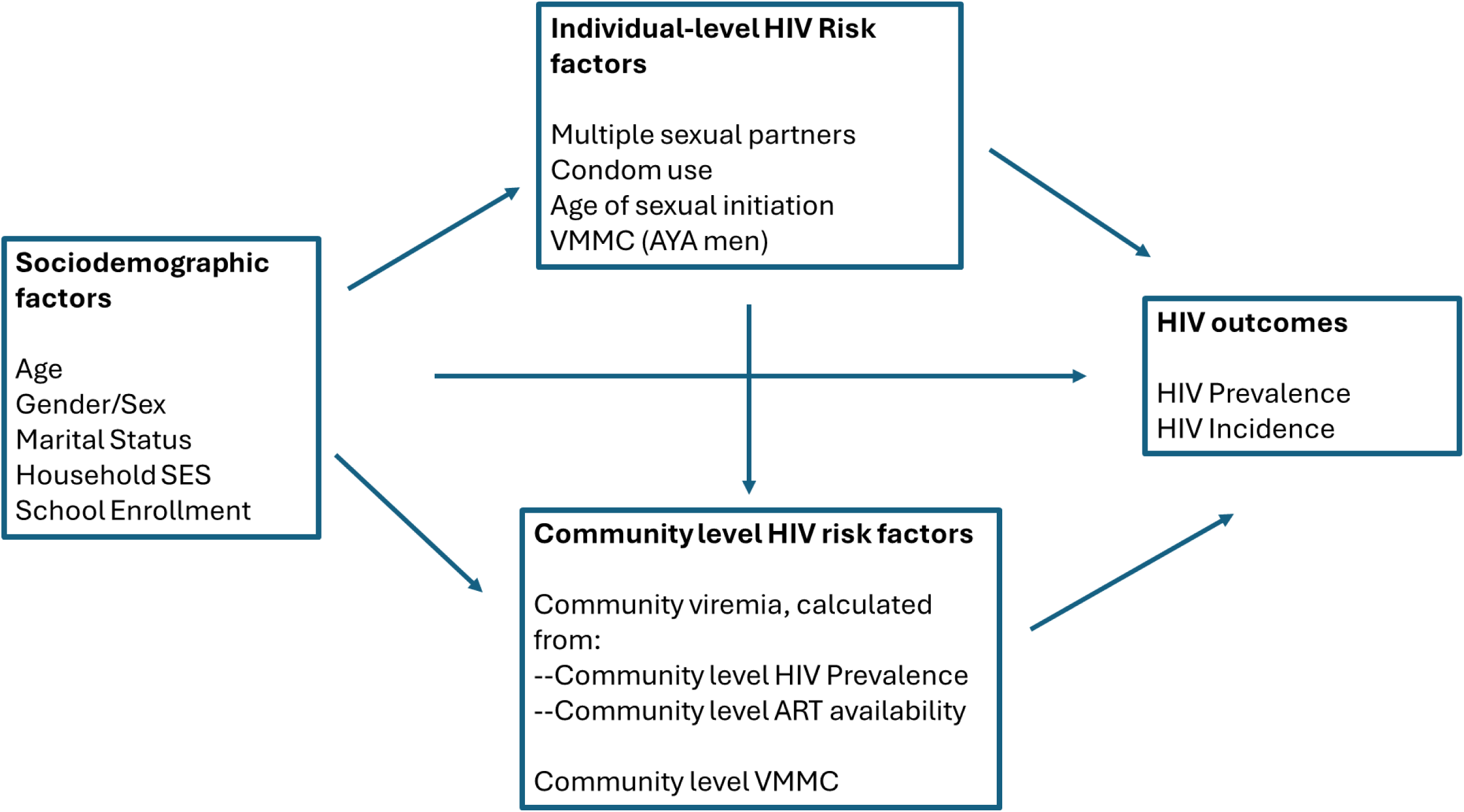
Conceptual Model.

**Supplementary Figure 2.**
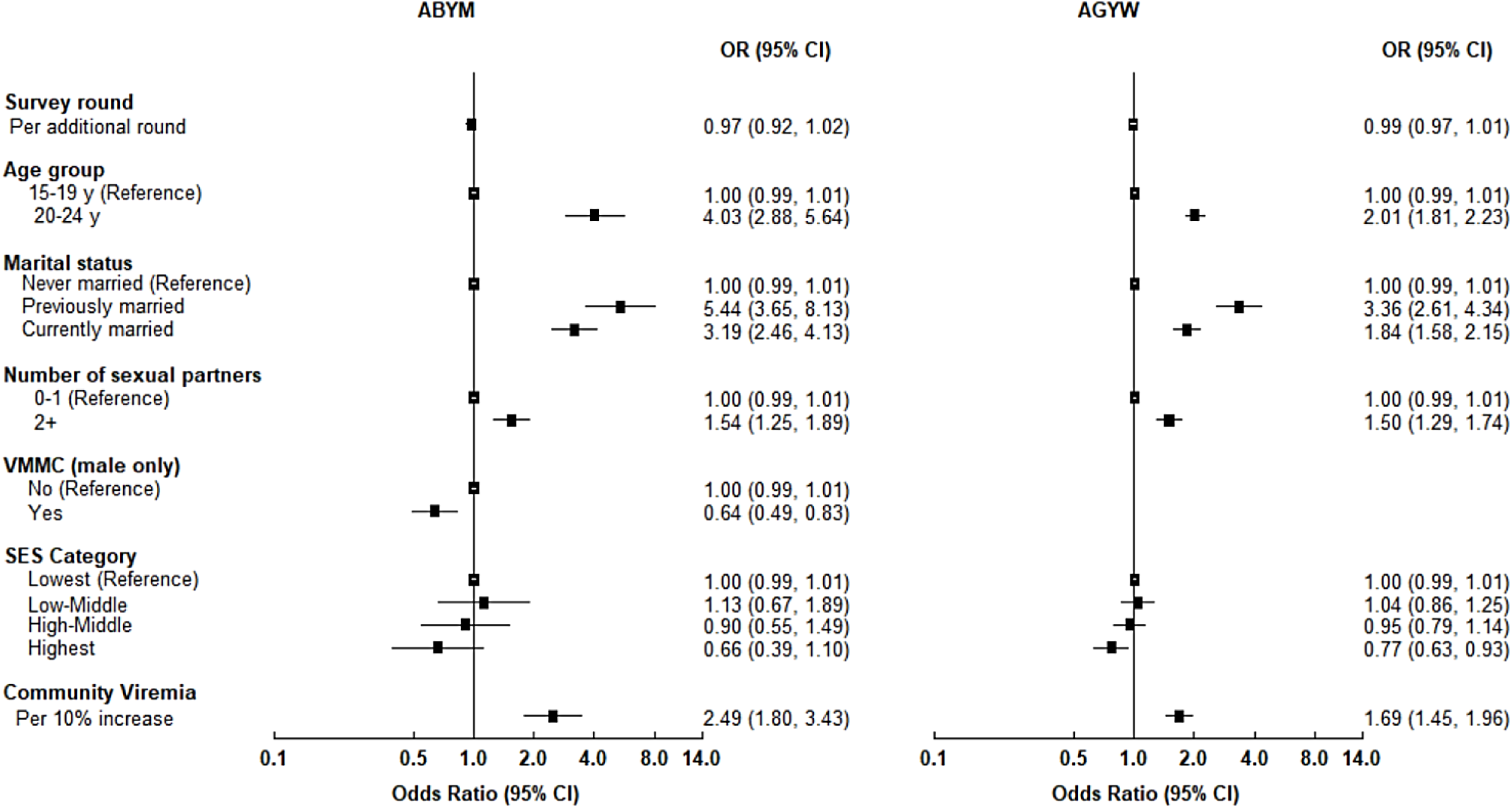
Univariate associations between selected individual-level risk factors and HIV prevalence, by gender. ABYM = Adolescent boys and young men, AGYW = Adolescent girls and young women, CI = Confidence intervals, OR= Odds Ratio, SES= Socio-economic status, VMMC = Voluntary Medical Male Circumcision. Community Viremia was calculated from (HIV Prevalence) - 0.9*(ART Coverage)*(HIV Prevalence). In models for ABYM, community viremia among AGYW aged 15-30 years was used; in models for AGYW, community viremia among ABYM aged 15-34 years was used. Number of sex partners refers to the last 12 months prior to the interview date.

**Supplementary Figure 3.**
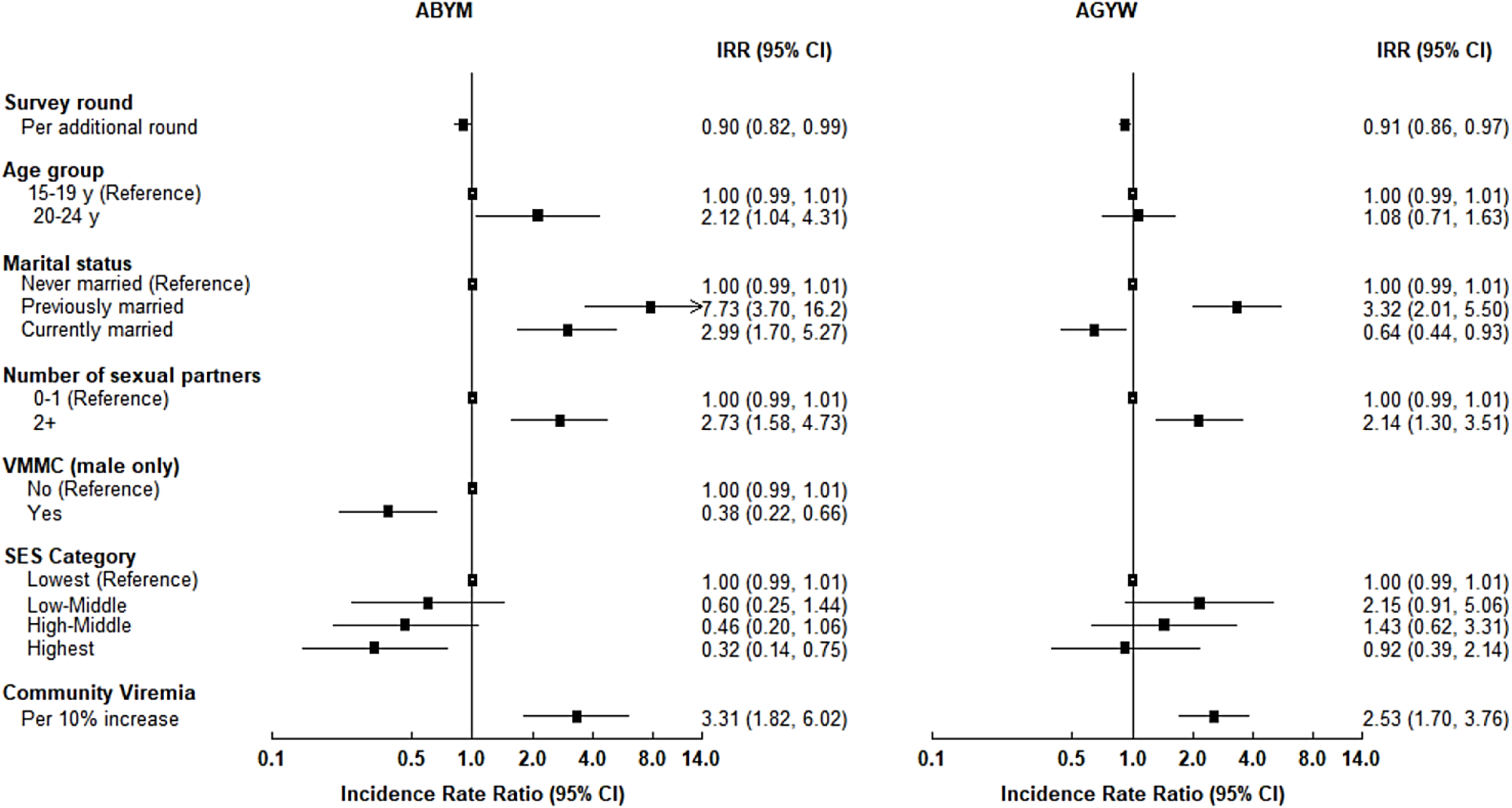
Univariate associations between selected individual-level risk factors and HIV incidence, by gender. ABYM = Adolescent boys and young men, AGYW = Adolescent girls and young women, CI = Confidence intervals, SES= Socio-economic status, VMMC = Voluntary Medical Male Circumcision. Community Viremia was calculated from (HIV Prevalence) - 0.9*(ART Coverage)*(HIV Prevalence). Associations with HIV prevalence are adjusted odds ratios. Associations with HIV incidence are adjusted incidence rate ratios. In models for ABYM, community viremia among AGYW aged 15-30 years was used; in models for AGYW, community viremia among ABYM aged 15-34 years was used. Number of sex partners refers to the last 12 months prior to the interview date.

**Supplemental Table 1.** HIV Prevalence and HIV Incidence Among Young Women and Young Men 15-24 Years, Rakai District, Uganda, 2005-2020.

|  |  |  | RCCS Round |  |  |  |  |  |  |  |  |  |  |
| --- | --- | --- | --- | --- | --- | --- | --- | --- | --- | --- | --- | --- | --- |
| Model Type | Gender | Age Group | N | 11 | 12 | 13 | 14 | 15 | 16 | 17 | 18 | 19 | P-value |
| Prevalence | Males | 15-24 | 22942 | 2.4 | 1.9 | 2.3 | 2.5 | 2.5 | 2.0 | 1.6 | 1.5 | 1.4 | 0.590 |
|  |  | 15-19 | 12338 | 0.0 | 0.7 | 0.8 | 1.1 | 1.3 | 0.4 | 0.4 | 0.9 | 1.1 | 0.017 |
|  |  | 20-24 | 10604 | 4.6 | 3.3 | 4.3 | 4.4 | 4.3 | 4.1 | 3.2 | 2.3 | 2.0 | 0.033 |
|  | Females | 15-24 | 29250 | 7.9 | 7.9 | 7.4 | 7.1 | 7.7 | 7.0 | 7.1 | 5.3 | 3.5 | 0.073 |
|  |  | 15-19 | 13935 | 4.6 | 2.6 | 2.2 | 2.5 | 3.3 | 2.3 | 3.3 | 1.9 | 1.3 | 0.002 |
|  |  | 20-24 | 15315 | 10.3 | 12.1 | 12.3 | 12.1 | 12.1 | 11.8 | 11.2 | 9.2 | 6.1 | 0.500 |
| Incidence | Males | 15-24 | 10668 | 4.2 | 6.8 | 6.4 | 8.9 | 4.5 | 2.1 | 2.5 | 2.8 | 3.0 | 0.008 |
|  |  | 15-19 | 4677 | 0.0 | 4.7 | 3.4 | 3.1 | 3.9 | 1.2 | 0.0 | 0.0 | 0.0 | 0.019 |
|  |  | 20-24 | 5991 | 6.3 | 8.0 | 8.6 | 13.6 | 5.1 | 2.9 | 4.4 | 5.2 | 5.1 | 0.095 |
|  | Females | 15-24 | 11761 | 14.2 | 14.1 | 10.9 | 13.8 | 10.2 | 12.7 | 12.1 | 2.1 | 4.9 | <0.001 |
|  |  | 15-19 | 4298 | 23.7 | 9.8 | 6.4 | 5.2 | 6.8 | 7.0 | 4.3 | 0.0 | 1.2 | <0.001 |
|  |  | 20-24 | 7463 | 10.5 | 16.0 | 13.0 | 20.0 | 12.6 | 15.5 | 17.3 | 3.6 | 7.7 | 0.085 |

*P-values for prevalence models were estimated using GEE (binary with logit link); p-values for incidence models were estimated using GLM Poisson regression, offset log(time)*
*HIV prevalence is percentage.*
*HIV incidence per 1000 person-year (no. of new HIV-positive)*

**Supplemental Table 2.**
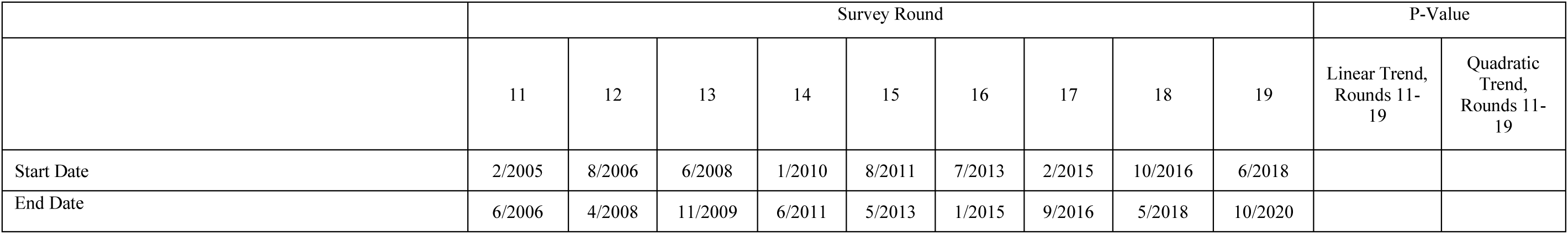
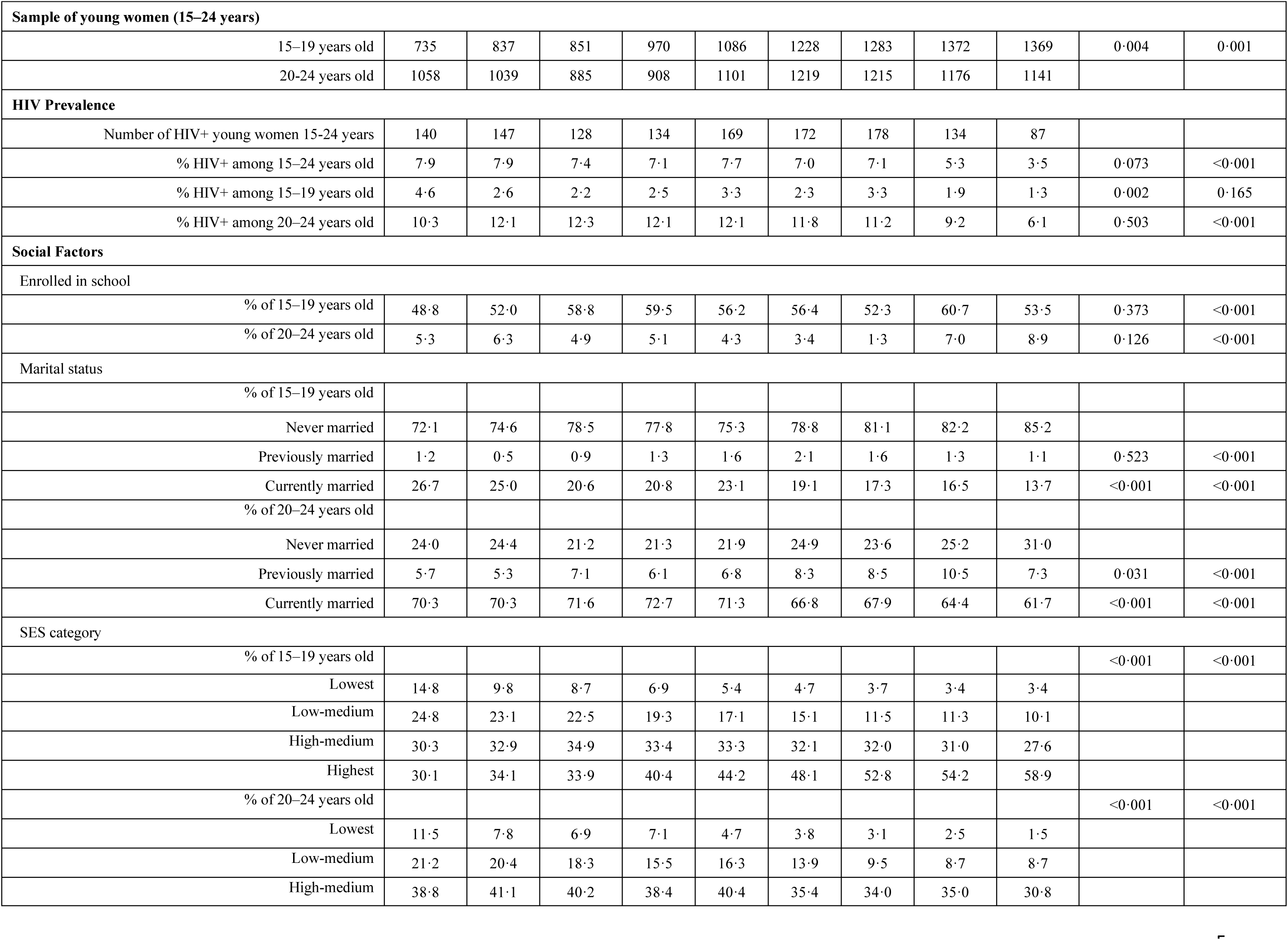
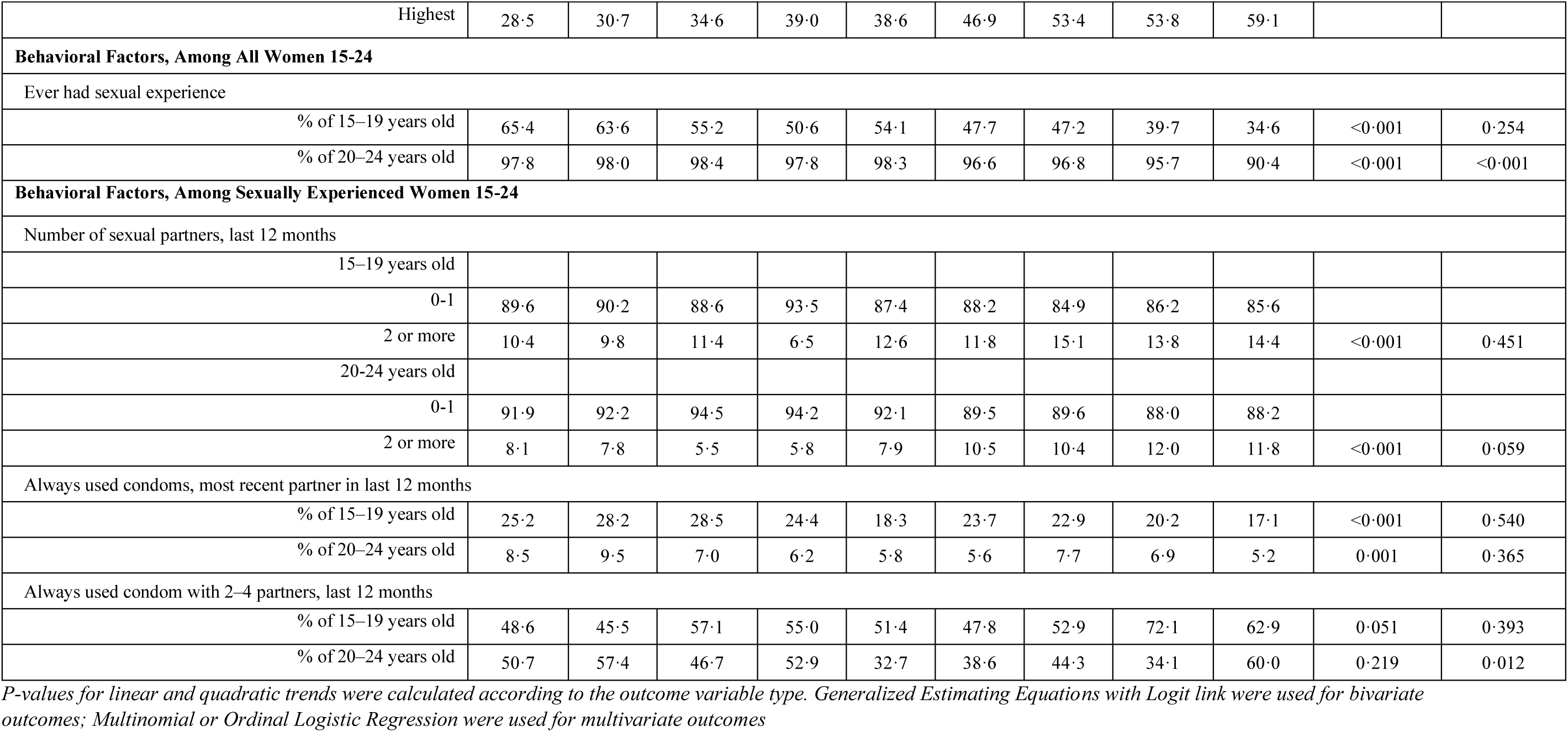
Trends in HIV Prevalence, Social Factors, and Behavioral Factors among Young Women 15–24 Years, Rakai District, Uganda, 2005-2020.

|  | Survey Round |  |  |  |  |  |  |  |  | P-Value |  |
| --- | --- | --- | --- | --- | --- | --- | --- | --- | --- | --- | --- |
|  | 11 | 12 | 13 | 14 | 15 | 16 | 17 | 18 | 19 | Linear Trend,<br>Rounds 11-19 | Quadratic Trend,<br>Rounds 11-19 |
| Start Date | 2/2005 | 8/2006 | 6/2008 | 1/2010 | 8/2011 | 7/2013 | 2/2015 | 10/2016 | 6/2018 |  |  |
| End Date | 6/2006 | 4/2008 | 11/2009 | 6/2011 | 5/2013 | 1/2015 | 9/2016 | 5/2018 | 10/2020 |  |  |
| **Sample of young women (15–24 years)** |  |  |  |  |  |  |  |  |  |  |  |
| 15–19 years old | 735 | 837 | 851 | 970 | 1086 | 1228 | 1283 | 1372 | 1369 | 0.004 | 0.001 |
| 20–24 years old | 1058 | 1039 | 885 | 908 | 1101 | 1219 | 1215 | 1176 | 1141 |  |  |
| **HIV Prevalence** |  |  |  |  |  |  |  |  |  |  |  |
| Number of HIV+ young women 15–24 years | 140 | 147 | 128 | 134 | 169 | 172 | 178 | 134 | 87 |  |  |
| % HIV+ among 15–24 years old | 7.9 | 7.9 | 7.4 | 7.1 | 7.7 | 7.0 | 7.1 | 5.3 | 3.5 | 0.073 | <0.001 |
| % HIV+ among 15–19 years old | 4.6 | 2.6 | 2.2 | 2.5 | 3.3 | 2.3 | 3.3 | 1.9 | 1.3 | 0.002 | 0.165 |
| % HIV+ among 20–24 years old | 10.3 | 12.1 | 12.3 | 12.1 | 12.1 | 11.8 | 11.2 | 9.2 | 6.1 | 0.503 | <0.001 |
| **Social Factors** |  |  |  |  |  |  |  |  |  |  |  |
| Enrolled in school |  |  |  |  |  |  |  |  |  |  |  |
| % of 15–19 years old | 48.8 | 52.0 | 58.8 | 59.5 | 56.2 | 56.4 | 52.3 | 60.7 | 53.5 | 0.373 | <0.001 |
| % of 20–24 years old | 5.3 | 6.3 | 4.9 | 5.1 | 4.3 | 3.4 | 1.3 | 7.0 | 8.9 | 0.126 | <0.001 |
| Marital status |  |  |  |  |  |  |  |  |  |  |  |
| % of 15–19 years old |  |  |  |  |  |  |  |  |  |  |  |
| Never married | 72.1 | 74.6 | 78.5 | 77.8 | 75.3 | 78.8 | 81.1 | 82.2 | 85.2 |  |  |
| Previously married | 1.2 | 0.5 | 0.9 | 1.3 | 1.6 | 2.1 | 1.6 | 1.3 | 1.1 | 0.523 | <0.001 |
| Currently married | 26.7 | 25.0 | 20.6 | 20.8 | 23.1 | 19.1 | 17.3 | 16.5 | 13.7 | <0.001 | <0.001 |
| % of 20–24 years old |  |  |  |  |  |  |  |  |  |  |  |
| Never married | 24.0 | 24.4 | 21.2 | 21.3 | 21.9 | 24.9 | 23.6 | 25.2 | 31.0 |  |  |
| Previously married | 5.7 | 5.3 | 7.1 | 6.1 | 6.8 | 8.3 | 8.5 | 10.5 | 7.3 | 0.031 | <0.001 |
| Currently married | 70.3 | 70.3 | 71.6 | 72.7 | 71.3 | 66.8 | 67.9 | 64.4 | 61.7 | <0.001 | <0.001 |
| SES category |  |  |  |  |  |  |  |  |  |  |  |
| % of 15–19 years old |  |  |  |  |  |  |  |  |  | <0.001 | <0.001 |
| Lowest | 14.8 | 9.8 | 8.7 | 6.9 | 5.4 | 4.7 | 3.7 | 3.4 | 3.4 |  |  |
| Low-medium | 24.8 | 23.1 | 22.5 | 19.3 | 17.1 | 15.1 | 11.5 | 11.3 | 10.1 |  |  |
| High-medium | 30.3 | 32.9 | 34.9 | 33.4 | 33.3 | 32.1 | 32.0 | 31.0 | 27.6 |  |  |
| Highest | 30.1 | 34.1 | 33.9 | 40.4 | 44.2 | 48.1 | 52.8 | 54.2 | 58.9 |  |  |
| % of 20–24 years old |  |  |  |  |  |  |  |  |  | <0.001 | <0.001 |
| Lowest | 11.5 | 7.8 | 6.9 | 7.1 | 4.7 | 3.8 | 3.1 | 2.5 | 1.5 |  |  |
| Low-medium | 21.2 | 20.4 | 18.3 | 15.5 | 16.3 | 13.9 | 9.5 | 8.7 | 8.7 |  |  |
| High-medium | 38.8 | 41.1 | 40.2 | 38.4 | 40.4 | 35.4 | 34.0 | 35.0 | 30.8 |  |  |
| Highest | 28.5 | 30.7 | 34.6 | 39.0 | 38.6 | 46.9 | 53.4 | 53.8 | 59.1 |  |  |
| **Behavioral Factors, Among All Women 15-24** |  |  |  |  |  |  |  |  |  |  |  |
| Ever had sexual experience |  |  |  |  |  |  |  |  |  |  |  |
| % of 15–19 years old | 65.4 | 63.6 | 55.2 | 50.6 | 54.1 | 47.7 | 47.2 | 39.7 | 34.6 | <0.001 | 0.254 |
| % of 20–24 years old | 97.8 | 98.0 | 98.4 | 97.8 | 98.3 | 96.6 | 96.8 | 95.7 | 90.4 | <0.001 | <0.001 |
| **Behavioral Factors, Among Sexually Experienced Women 15-24** |  |  |  |  |  |  |  |  |  |  |  |
| Number of sexual partners, last 12 months |  |  |  |  |  |  |  |  |  |  |  |
| 15–19 years old |  |  |  |  |  |  |  |  |  |  |  |
| 0-1 | 89.6 | 90.2 | 88.6 | 93.5 | 87.4 | 88.2 | 84.9 | 86.2 | 85.6 |  |  |
| 2 or more | 10.4 | 9.8 | 11.4 | 6.5 | 12.6 | 11.8 | 15.1 | 13.8 | 14.4 | <0.001 | 0.451 |
| 20-24 years old |  |  |  |  |  |  |  |  |  |  |  |
| 0-1 | 91.9 | 92.2 | 94.5 | 94.2 | 92.1 | 89.5 | 89.6 | 88.0 | 88.2 |  |  |
| 2 or more | 8.1 | 7.8 | 5.5 | 5.8 | 7.9 | 10.5 | 10.4 | 12.0 | 11.8 | <0.001 | 0.059 |
| Always used condoms, most recent partner in last 12 months |  |  |  |  |  |  |  |  |  |  |  |
| % of 15–19 years old | 25.2 | 28.2 | 28.5 | 24.4 | 18.3 | 23.7 | 22.9 | 20.2 | 17.1 | <0.001 | 0.540 |
| % of 20–24 years old | 8.5 | 9.5 | 7.0 | 6.2 | 5.8 | 5.6 | 7.7 | 6.9 | 5.2 | 0.001 | 0.365 |
| Always used condom with 2–4 partners, last 12 months |  |  |  |  |  |  |  |  |  |  |  |
| % of 15–19 years old | 48.6 | 45.5 | 57.1 | 55.0 | 51.4 | 47.8 | 52.9 | 72.1 | 62.9 | 0.051 | 0.393 |
| % of 20–24 years old | 50.7 | 57.4 | 46.7 | 52.9 | 32.7 | 38.6 | 44.3 | 34.1 | 60.0 | 0.219 | 0.012 |

*P-values for linear and quadratic trends were calculated according to the outcome variable type. Generalized Estimating Equations with Logit link were used for bivariate outcomes; Multinomial or Ordinal Logistic Regression were used for multivariate outcomes*

**Supplemental Table 3.**
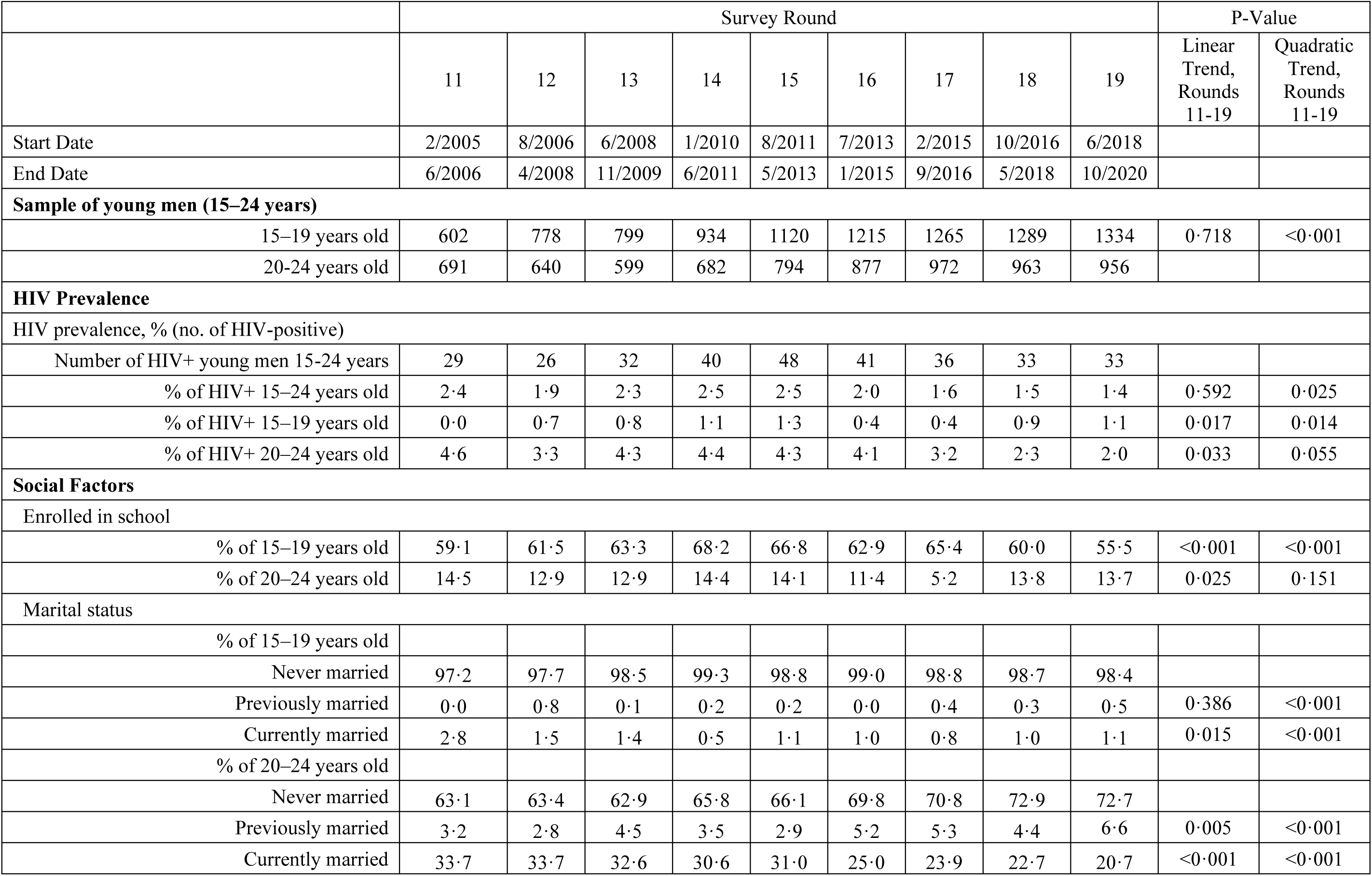
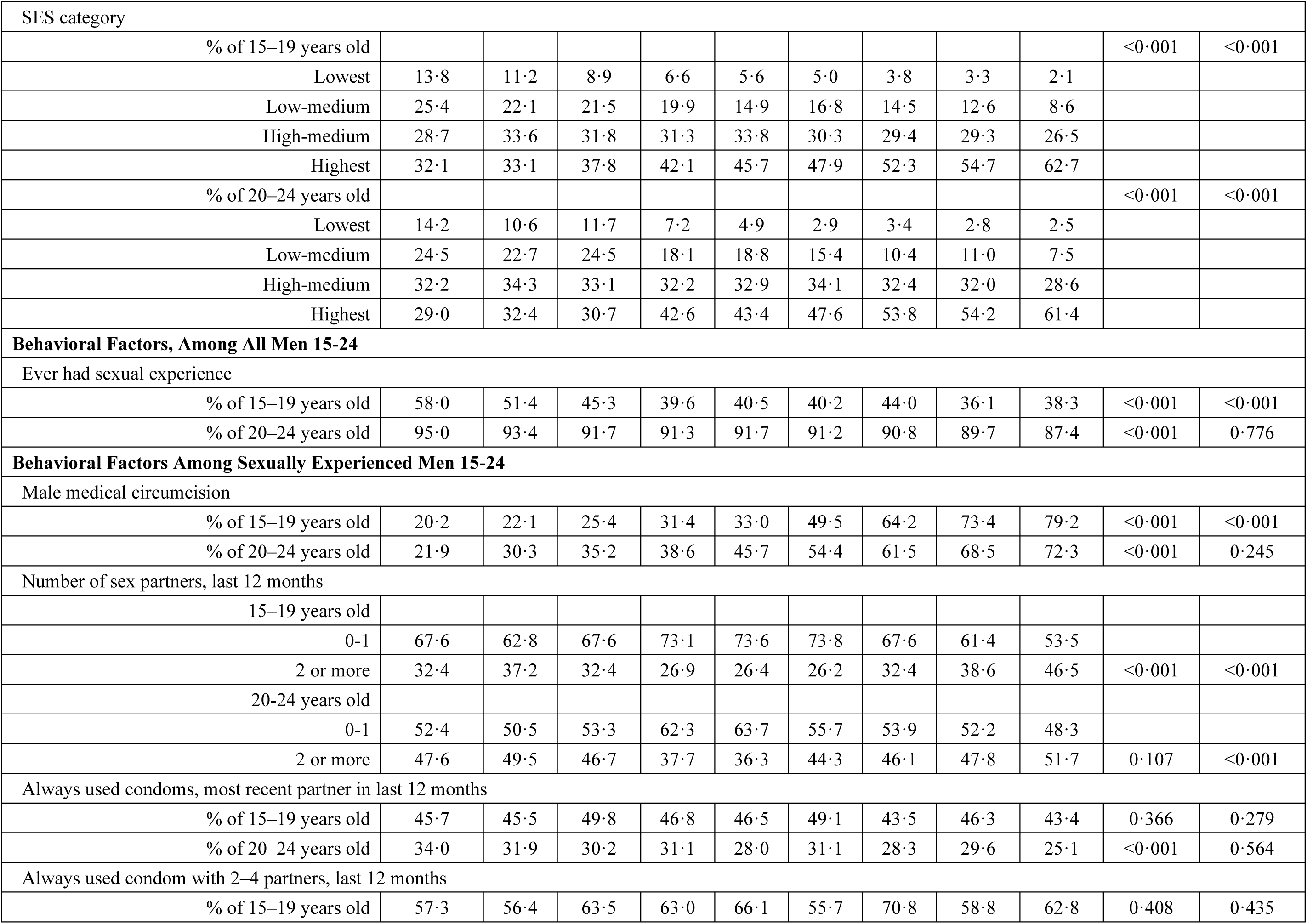
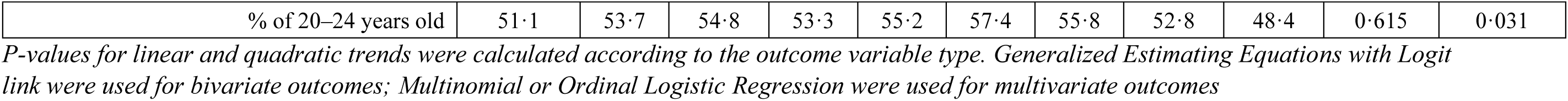
Trends in HIV Prevalence, Social Factors, and HIV Risk Factors in Young Men 15–24 Years in 30 Communities, Rakai District, Uganda, 2005-2020.

|  | Survey Round |  |  |  |  |  |  |  |  | P-Value |  |
| --- | --- | --- | --- | --- | --- | --- | --- | --- | --- | --- | --- |
|  | 11 | 12 | 13 | 14 | 15 | 16 | 17 | 18 | 19 | Linear Trend, Rounds 11-19 | Quadratic Trend, Rounds 11-19 |
| Start Date | 2/2005 | 8/2006 | 6/2008 | 1/2010 | 8/2011 | 7/2013 | 2/2015 | 10/2016 | 6/2018 |  |  |
| End Date | 6/2006 | 4/2008 | 11/2009 | 6/2011 | 5/2013 | 1/2015 | 9/2016 | 5/2018 | 10/2020 |  |  |
| **Sample of young men (15–24 years)** |  |  |  |  |  |  |  |  |  |  |  |
| 15–19 years old | 602 | 778 | 799 | 934 | 1120 | 1215 | 1265 | 1289 | 1334 | 0.718 | <0.001 |
| 20-24 years old | 691 | 640 | 599 | 682 | 794 | 877 | 972 | 963 | 956 |  |  |
| **HIV Prevalence** |  |  |  |  |  |  |  |  |  |  |  |
| HIV prevalence, % (no. of HIV-positive) |  |  |  |  |  |  |  |  |  |  |  |
| Number of HIV+ young men 15-24 years | 29 | 26 | 32 | 40 | 48 | 41 | 36 | 33 | 33 |  |  |
| % of HIV+ 15–24 years old | 2.4 | 1.9 | 2.3 | 2.5 | 2.5 | 2.0 | 1.6 | 1.5 | 1.4 | 0.592 | 0.025 |
| % of HIV+ 15–19 years old | 0.0 | 0.7 | 0.8 | 1.1 | 1.3 | 0.4 | 0.4 | 0.9 | 1.1 | 0.017 | 0.014 |
| % of HIV+ 20–24 years old | 4.6 | 3.3 | 4.3 | 4.4 | 4.3 | 4.1 | 3.2 | 2.3 | 2.0 | 0.033 | 0.055 |
| **Social Factors** |  |  |  |  |  |  |  |  |  |  |  |
| Enrolled in school |  |  |  |  |  |  |  |  |  |  |  |
| % of 15–19 years old | 59.1 | 61.5 | 63.3 | 68.2 | 66.8 | 62.9 | 65.4 | 60.0 | 55.5 | <0.001 | <0.001 |
| % of 20–24 years old | 14.5 | 12.9 | 12.9 | 14.4 | 14.1 | 11.4 | 5.2 | 13.8 | 13.7 | 0.025 | 0.151 |
| Marital status |  |  |  |  |  |  |  |  |  |  |  |
| % of 15–19 years old |  |  |  |  |  |  |  |  |  |  |  |
| Never married | 97.2 | 97.7 | 98.5 | 99.3 | 98.8 | 99.0 | 98.8 | 98.7 | 98.4 |  |  |
| Previously married | 0.0 | 0.8 | 0.1 | 0.2 | 0.2 | 0.0 | 0.4 | 0.3 | 0.5 | 0.386 | <0.001 |
| Currently married | 2.8 | 1.5 | 1.4 | 0.5 | 1.1 | 1.0 | 0.8 | 1.0 | 1.1 | 0.015 | <0.001 |
| % of 20–24 years old |  |  |  |  |  |  |  |  |  |  |  |
| Never married | 63.1 | 63.4 | 62.9 | 65.8 | 66.1 | 69.8 | 70.8 | 72.9 | 72.7 |  |  |
| Previously married | 3.2 | 2.8 | 4.5 | 3.5 | 2.9 | 5.2 | 5.3 | 4.4 | 6.6 | 0.005 | <0.001 |
| Currently married | 33.7 | 33.7 | 32.6 | 30.6 | 31.0 | 25.0 | 23.9 | 22.7 | 20.7 | <0.001 | <0.001 |

| SES category |  |  |  |  |  |  |  |  |  |  |  |
| --- | --- | --- | --- | --- | --- | --- | --- | --- | --- | --- | --- |
| % of 15–19 years old |  |  |  |  |  |  |  |  |  | <0.001 | <0.001 |
| Lowest | 13.8 | 11.2 | 8.9 | 6.6 | 5.6 | 5.0 | 3.8 | 3.3 | 2.1 |  |  |
| Low-medium | 25.4 | 22.1 | 21.5 | 19.9 | 14.9 | 16.8 | 14.5 | 12.6 | 8.6 |  |  |
| High-medium | 28.7 | 33.6 | 31.8 | 31.3 | 33.8 | 30.3 | 29.4 | 29.3 | 26.5 |  |  |
| Highest | 32.1 | 33.1 | 37.8 | 42.1 | 45.7 | 47.9 | 52.3 | 54.7 | 62.7 |  |  |
| % of 20–24 years old |  |  |  |  |  |  |  |  |  | <0.001 | <0.001 |
| Lowest | 14.2 | 10.6 | 11.7 | 7.2 | 4.9 | 2.9 | 3.4 | 2.8 | 2.5 |  |  |
| Low-medium | 24.5 | 22.7 | 24.5 | 18.1 | 18.8 | 15.4 | 10.4 | 11.0 | 7.5 |  |  |
| High-medium | 32.2 | 34.3 | 33.1 | 32.2 | 32.9 | 34.1 | 32.4 | 32.0 | 28.6 |  |  |
| Highest | 29.0 | 32.4 | 30.7 | 42.6 | 43.4 | 47.6 | 53.8 | 54.2 | 61.4 |  |  |
| **Behavioral Factors, Among All Men 15-24** |  |  |  |  |  |  |  |  |  |  |  |
| Ever had sexual experience |  |  |  |  |  |  |  |  |  |  |  |
| % of 15–19 years old | 58.0 | 51.4 | 45.3 | 39.6 | 40.5 | 40.2 | 44.0 | 36.1 | 38.3 | <0.001 | <0.001 |
| % of 20–24 years old | 95.0 | 93.4 | 91.7 | 91.3 | 91.7 | 91.2 | 90.8 | 89.7 | 87.4 | <0.001 | 0.776 |
| **Behavioral Factors Among Sexually Experienced Men 15-24** |  |  |  |  |  |  |  |  |  |  |  |
| Male medical circumcision |  |  |  |  |  |  |  |  |  |  |  |
| % of 15–19 years old | 20.2 | 22.1 | 25.4 | 31.4 | 33.0 | 49.5 | 64.2 | 73.4 | 79.2 | <0.001 | <0.001 |
| % of 20–24 years old | 21.9 | 30.3 | 35.2 | 38.6 | 45.7 | 54.4 | 61.5 | 68.5 | 72.3 | <0.001 | 0.245 |
| Number of sex partners, last 12 months |  |  |  |  |  |  |  |  |  |  |  |
| 15–19 years old |  |  |  |  |  |  |  |  |  |  |  |
| 0-1 | 67.6 | 62.8 | 67.6 | 73.1 | 73.6 | 73.8 | 67.6 | 61.4 | 53.5 |  |  |
| 2 or more | 32.4 | 37.2 | 32.4 | 26.9 | 26.4 | 26.2 | 32.4 | 38.6 | 46.5 | <0.001 | <0.001 |
| 20-24 years old |  |  |  |  |  |  |  |  |  |  |  |
| 0-1 | 52.4 | 50.5 | 53.3 | 62.3 | 63.7 | 55.7 | 53.9 | 52.2 | 48.3 |  |  |
| 2 or more | 47.6 | 49.5 | 46.7 | 37.7 | 36.3 | 44.3 | 46.1 | 47.8 | 51.7 | 0.107 | <0.001 |
| Always used condoms, most recent partner in last 12 months |  |  |  |  |  |  |  |  |  |  |  |
| % of 15–19 years old | 45.7 | 45.5 | 49.8 | 46.8 | 46.5 | 49.1 | 43.5 | 46.3 | 43.4 | 0.366 | 0.279 |
| % of 20–24 years old | 34.0 | 31.9 | 30.2 | 31.1 | 28.0 | 31.1 | 28.3 | 29.6 | 25.1 | <0.001 | 0.564 |
| Always used condom with 2–4 partners, last 12 months |  |  |  |  |  |  |  |  |  |  |  |
| % of 15–19 years old | 57.3 | 56.4 | 63.5 | 63.0 | 66.1 | 55.7 | 70.8 | 58.8 | 62.8 | 0.408 | 0.435 |
| % of 20–24 years old | 51·1 | 53·7 | 54·8 | 53·3 | 55·2 | 57·4 | 55·8 | 52·8 | 48·4 | 0·615 | 0·031 |

*P-values for linear and quadratic trends were calculated according to the outcome variable type. Generalized Estimating Equations with Logit link were used for bivariate outcomes; Multinomial or Ordinal Logistic Regression were used for multivariate outcomes*

**Supplemental Table 4.** Trends in Community Level HIV Risk Factors in 30 Communities, Rakai District, Uganda, 2005-2020.

|  |  |  |  |  |  |  |  |  |  |  | P-Value |  |
| --- | --- | --- | --- | --- | --- | --- | --- | --- | --- | --- | --- | --- |
|  |  |  |  |  |  |  |  |  |  |  | Linear Trend | Quadratic Trend |
| Women | 11 | 12 | 13 | 14 | 15 | 16 | 17 | 18 | 19 |  |  |  |
| Community HIV Prevalence | 11.9 | 11.4 | 11.8 | 11.6 | 12.3 | 11.4 | 11.4 | 9.5 | 8.2 |  | <0.001 | 0.007 |
| % ART among people living with HIV | 6.5 | 5.9 | 9.8 | 11.5 | 18.4 | 46.9 | 62.7 | 67.6 | 72.8 |  | <0.001 | <0.001 |
| Community Viremia | 11.2 | 10.7 | 10.8 | 10.3 | 10.1 | 6.7 | 5.1 | 3.8 | 2.9 |  | <0.001 | 0.001 |
|  |  |  |  |  |  |  |  |  |  |  | P-Value |  |
|  |  |  |  |  |  |  |  |  |  |  | Linear Trend | Quadratic Trend |
| Men | 11 | 12 | 13 | 14 | 15 | 16 | 17 | 18 | 19 |  |  |  |
| Community HIV Prevalence | 8.1 | 7.0 | 8.4 | 7.6 | 7.4 | 6.6 | 5.8 | 5.1 | 4.2 |  | <0.001 | 0.040 |
| % ART among people living with HIV | 4.0 | 5.9 | 9.5 | 10.5 | 19.1 | 34.7 | 50.0 | 55.8 | 65.8 |  | <0.001 | <0.001 |
| Community Viremia | 8.0 | 6.9 | 7.9 | 7.1 | 6.1 | 4.6 | 3.3 | 2.6 | 1.8 |  | <0.001 | 0.230 |
| Community Prevalence Male Medical Circumcision | 21.8 | 29.7 | 34.6 | 39.6 | 42.8 | 55.8 | 62.8 | 70.2 | 73.8 |  | <0.001 | 0.029 |

*Community level variables are calculated among men ages 15-34*
*P-values for linear and quadratic trends were calculated using Generalized Estimating Equations with Gaussian link*

**Supplemental Table 5.** Predictors of HIV Prevalence Among Young Adults, Rakai District, Uganda, Ages 15-24, 2005-2020.

|  | Men |  |  |  |  | Women |  |  |  |  |
| --- | --- | --- | --- | --- | --- | --- | --- | --- | --- | --- |
|  | N | OR | CI Low | CI High | P-value | N | OR | CI Low | CI High | P-value |
| *Individual Level Variables* |  |  |  |  |  |  |  |  |  |  |
| Survey Round | 8508 | 1.06 | 0.99 | 1.13 | 0.1201 | 12963 | 1.00 | 0.98 | 1.03 | 0.8081 |
| Age Group |  |  |  |  |  |  |  |  |  |  |
| 15-19 | 2798 | 1.00 |  |  |  | 4087 | 1.00 |  |  |  |
| 20-24 | 5710 | 2.84 | 1.90 | 4.25 | <0.0001 | 8876 | 1.82 | 1.63 | 2.04 | <0.0001 |
| Marriage Status |  |  |  |  |  |  |  |  |  |  |
| Never Married | 6168 | 1.00 |  |  |  | 3765 | 1.00 |  |  |  |
| Previously Married | 312 | 4.08 | 2.70 | 6.17 | <0.0001 | 751 | 2.55 | 1.97 | 3.28 | <0.0001 |
| Currently Married | 2028 | 2.36 | 1.79 | 3.10 | <0.0001 | 8447 | 1.57 | 1.33 | 1.85 | <0.0001 |
| Number of Sex Partners in Past 12 Months |  |  |  |  |  |  |  |  |  |  |
| 0-1 | 4944 | 1.00 |  |  |  | 11680 | 1.00 |  |  |  |
| 2 or More | 3564 | 1.39 | 1.12 | 1.73 | 0.0028 | 1283 | 1.52 | 1.30 | 1.78 | <0.0001 |
| SES Category |  |  |  |  |  |  |  |  |  |  |
| Lowest | 533 | 1.00 |  |  |  | 774 | 1.00 |  |  |  |
| Low-Middle | 1449 | 1.07 | 0.66 | 1.73 | 0.7798 | 2047 | 1.04 | 0.86 | 1.27 | 0.6734 |
| High-Middle | 2806 | 0.83 | 0.52 | 1.34 | 0.4550 | 4711 | 0.94 | 0.77 | 1.13 | 0.4925 |
| Highest | 3720 | 0.62 | 0.38 | 1.01 | 0.0546 | 5431 | 0.78 | 0.64 | 0.95 | 0.0163 |
| Male Medical Circumcision |  |  |  |  |  |  |  |  |  |  |
| No | 4234 | 1.00 |  |  |  |  |  |  |  |  |
| Yes | 4274 | 0.64 | 0.48 | 0.86 | 0.0026 |  |  |  |  |  |
| *Community Level Variables* |  |  |  |  |  |  |  |  |  |  |
| Community Viremia |  | 1.06 | 1.02 | 1.09 | 0.0027 |  | 1.03 | 1.02 | 1.05 | <0.0001 |

*Community Viremia is calculated from (HIV Prevalence) - 0.9\*(ART Coverage)\*(HIV Prevalence)*
*In models for men, we use community viremia among women 15-30; in models for women, we use community viremia among men 15-34. These ranges were selected as they are the most likely groups to transmit HIV to AYA. AYA women tend to have somewhat older male partners up to approximately age 34, while AYA men have slightly younger female partners up to approximately age 30.*

**Supplemental Table 6.** Predictors of HIV Incidence Among Young Adults, Rakai District, Uganda, Ages 15-24, 2005-2020.

|  | Men |  |  |  |  | Women |  |  |  |  |
| --- | --- | --- | --- | --- | --- | --- | --- | --- | --- | --- |
|  | *N* | *IRR* | *CI Low* | *CI High* | *P-value* | *N* | *IRR* | *CI Low* | *CI High* | *P-value* |
| *Individual Level Variables* |  |  |  |  |  |  |  |  |  |  |
| Survey Round | 4877 | 1.05 | 0.93 | 1.18 | 0.4590 | 5767 | 0.94 | 0.87 | 1.03 | 0.1725 |
| Age Group |  |  |  |  |  |  |  |  |  |  |
| 15-19 | 1431 | 1.00 |  |  |  | 1335 | 1.00 |  |  |  |
| 20-24 | 3446 | 1.15 | 0.50 | 2.65 | 0.7472 | 4432 | 1.31 | 0.80 | 2.13 | 0.2786 |
| Marriage Status |  |  |  |  |  |  |  |  |  |  |
| Never Married | 3561 | 1.00 |  |  |  | 1745 | 1.00 |  |  |  |
| Previously Married | 178 | 5.89 | 2.66 | 13.04 | <0.0001 | 231 | 2.76 | 1.58 | 4.80 | 0.0003 |
| Currently Married | 1138 | 2.63 | 1.40 | 4.95 | 0.0027 | 3791 | 0.58 | 0.37 | 0.90 | 0.0144 |
| Number of Sex Partners in Past 12 Months |  |  |  |  |  |  |  |  |  |  |
| 0-1 | 2809 | 1.00 |  |  |  | 5390 | 1.00 |  |  |  |
| 2 or More | 2068 | 2.41 | 1.36 | 4.27 | 0.0025 | 377 | 1.76 | 1.04 | 2.97 | 0.0356 |
| SES Category |  |  |  |  |  |  |  |  |  |  |
| Lowest | 319 |  |  |  |  | 353 | 1.00 |  |  |  |
| Low-Middle | 865 | 0.66 | 0.27 | 1.62 | 0.3604 | 974 | 2.23 | 0.94 | 5.28 | 0.0677 |
| High-Middle | 1605 | 0.59 | 0.25 | 1.39 | 0.2266 | 2003 | 1.54 | 0.66 | 3.62 | 0.3213 |
| Highest | 2088 | 0.45 | 0.19 | 1.08 | 0.0750 | 2437 | 1.13 | 0.47 | 2.74 | 0.7844 |
| Male Medical Circumcision |  |  |  |  |  |  |  |  |  |  |
| No | 2300 | 1.00 |  |  |  |  |  |  |  |  |
| Yes | 2577 | 0.42 | 0.23 | 0.76 | 0.0046 |  |  |  |  |  |
| *Community Level Variables* |  |  |  |  |  |  |  |  |  |  |
| Community Viremia |  | 1.06 | 0.99 | 1.14 | 0.0967 |  | 1.05 | 1.00 | 1.11 | 0.0387 |

*Community Viremia is calculated from (HIV Prevalence) - 0.9\*(ART Coverage)\*(HIV Prevalence)*
*In models for men, we use community viremia among women 15-30; in models for women, we use community viremia among men 15-34. These ranges were selected as they are the most likely groups to transmit HIV to AYA. AYA women tend to have somewhat older male partners up to approximately age 34, while AYA men have slightly younger female partners up to approximately age 30.*

